# Mental health among pregnant women during the pandemic in Sweden– a mixed methods approach using data from the Mom2B mobile application for research

**DOI:** 10.1101/2020.12.18.20248466

**Authors:** Emma Fransson, Maria Karalexi, Mary Kimmel, Emma Bränn, Natasa Kollia, Vera van Zoest, Eira Nordling, Fotios C Papadopoulos, Alkistis Skalkidou

**Author notes:** **Correspondence to:** Alkistis Skalkidou, MD, PhD, Professor of Obstetrics and Gynecology, Department of Women’s and Children’s Health, Uppsala University, Sweden.

## Abstract

Public health emergencies such as the coronavirus (SARS-CoV-2) pandemic have significant impact on mental health, and have been shown to impact on already prevalent affective disorders during and after pregnancy. The aim of this study was to utilize modern tools to assess depressive and anxiety symptoms, as well as wellbeing and life changes in pregnant women during the pandemic in Sweden, where no lockdown has been in place.

Data from the Mom2B, a national ongoing mobile application-based study of pregnant and newly-delivered women were utilized. Participants (n= 1345) filled out self-report screeners of depression, anxiety and wellbeing. Questions about COVID symptoms and effects on life and health care were added from March 2020. Movement data was collected using the phone’s GPS sensor. Mood scores were compared with throughout the months of 2020 and to the levels of a previous collected material. Highest levels of depression and anxiety were evident in April and October 2020. Symptoms were higher among those feeling socially isolated, but not for those infected or with symptomatic family members. Wellbeing and mobility were strongly positively correlated and were lowest in April. Women reported on cancelled healthcare appointments and worry about their partners being absent from the delivery.

The Mom2B application enabled gathering information at a national level in real-time as the pandemic has been evolving. Levels of perinatal affective symptoms and low wellbeing were elevated compared with previous years as well as with months with fewer cases of SARS-Cov-2. Similar applications can help healthcare providers and governmental bodies to in real time monitor high-risk groups during crises, as well as to adjust measures and the support offered.

**Funding:** This project was funded by the Uppsala Region to AS, the Swedish Association of Local Authorities and Regions (SKR) to the department of Obstetrics and Gynecology, Akademiska University Hospital, the Swedish Research Council (Grant number 2020-01965) to AS, as well as the Fredrik and Inger Thuring’s Foundation to EF.

## Introduction

The coronavirus disease 2019 (COVID-19) has led to a huge healthcare crisis with significant sequelae, spreading to nearly all countries; the global death toll exceeds 1 million and the number of confirmed cases more than 50 million people (1). Beyond other adverse effects, the disease has posed significant threats in mental health with recent studies reporting an increase by 15-30% in anxiety and depression globally, whereas the infection itself may also affect the nervous system, and cause symptoms of mental illness (2, 3). Notably, patients with a recent mental disorder diagnosis show increased risk for COVID□19 infection (4). Importantly, the mental health effects of COVID-19 are expected to persist even after a vaccination is established, because of the neurologic sequelae of the disease, trauma of illness, loss of loved ones, and increased stress due to financial restraints.

Especially among pregnant women, recent studies in the US, China, Japan and Turkey have shown an increase by 25-50% in peripartum anxiety and depressive symptoms since the beginning of the pandemic (2, 5-7). Factors that may underlie these associations include the worry for the health of the pregnancy and the forthcoming infant, worry for the pandemic situation in general, economic consequences, grief for victims, misinformation about the virus, worry about not getting healthcare services as planned, social distancing/social isolation and travel restrictions (2, 3, 5). The epidemic has greatly affected hospital organization and procedures, especially those concerning the conditions and experiences of childbirth (8). Depressive and anxiety symptoms during pregnancy can increase the risk of postpartum depression; of note is that depression and anxiety in both pregnancy and postpartum have far-reaching effects including severe consequences on the mother’s future morbidity and the fetus’s psycho-emotional development (9-12). Young age, low maternal education, low income, unemployment and lack of partner support seem to be additional factors associated with higher risk of adverse perinatal mental health effects (13, 14).

In Sweden, with 115.000 births per year, medical interventions to address the risks associated with pregnancy and birth have been largely successful so far and have resulted in one of the lowest levels of maternal and neonatal morbidity and mortality worldwide (15). Sweden also offers a unique setting with as many as 92% of the Swedish population above the age of 12 owning a smartphone in 2019. Moreover, Sweden is one of the countries that did not implement strict lockdown measures, but relied on voluntary social distancing guidelines, including working from home when possible, reducing travel and social contacts and avoiding public transport (16). During the spring of 2020, no widespread public testing was in place in Sweden, and only very few non-hospitalized citizens were tested for COVID-19. Deaths peaked in late April, whereas the second wave during fall is ongoing with cases starting to surge again in October. The Swedish approach not to implement strict lockdown measures has garnered debate and may have resulted in greater or less anxiety, particularly for pregnant women.

This background of nearly free maternity care, low levels of maternal and neonatal morbidity, relative economic stability, in combination with the absence of bias from lockdown measures sequelae, provide a unique setting to study the impact related specifically to COVID-19 on perinatal depression and anxiety. We utilized data from a unique cohort of pregnant women recruited using the novel Mom2B mobile application for research, with the aim to examine: (1) if measures of mental health differed between the months of the pandemic as well as comparing with previous years, (2) if confirmed or possible infection and/or self-reported impact from the pandemic were associated with mental health measures, using validated instruments, (3) if mobility and corona-related internet searches correlated with symptoms and finally (4) which concerns were noted by Swedish pregnant women about their health care and wellbeing in relation to the pandemic.

## Methods

### Study population

The Mom2B cohort (www.mom2b.se) is a national ongoing mobile application-based mother cohort, introduced at the end of November 2019 to the App Store and Google Play. All Swedish-speaking women above 18 years of age owning a mobile smartphone who are either pregnant or have delivered within three months are eligible for participation. Information about the study is being posted in social media, as well as by using posters and brochures at local maternity clinics. By the end of October 2020, 1608 women were enrolled in the study and 1345 had answered at least one question related to mental health measures during pregnancy. The included self-report instruments were based on the results from a previous study in Uppsala County, Sweden (17), whereas the mobile application further recorded digital phenotyping data, such as movement patterns, internet and mobile use, as well as voice recordings, after receiving informed consent from participants. Privacy protection was central in the design of the application and data was collected only on i.e. relative geographical movement patterns, and not the exact position of participant. The application was a further development of the Beiwe research platform from the Harvard School of Public Health, adjusted to the Mom2B study questions and adhering to the General Data Protection Regulation (GDPR) regulations in Sweden (18).

### Comparison population

To compare the population in the Mom2B study to pre-pandemic years, we used data from 4879 pregnant women recruited in the Biologic Affect Stress Imagine and Cognition (BASIC)-study (17). BASIC was a population-based longitudinal cohort study, conducted at Uppsala University hospital in 2009-2019, with the aim to study biological and psychosocial aspects of perinatal depression. The participants, constituting around 21% of the background pregnant population, answered web-based self-report instruments regarding mental health twice during pregnancy and three times during postpartum. Women with valid outcome data from at least one-time during pregnancy were included in the analysis.

Ethical approval for the BASIC study was granted by the Regional Ethical Review Board of Uppsala, Sweden (Dnr 2009/171), and for the Mom2B-study by the Swedish Ethical Review Authority (Dnr 2019-01170), with amendments.

### Variables of interest

Perinatal anxiety and depressive symptoms were assessed with the Edinburgh Postnatal Depression Scale (EPDS)(19) a validated ten-item self-report questionnaire assessing depressive symptoms in the perinatal period with good psychometric properties (20). For the present study, a cutoff point of 13 points was used to define probable cases of depression during pregnancy, according to Swedish validation (21). Three items were used for measuring symptoms of anxiety (EPDS-3A)(22) with a cut-off score of six (22). EPDS was collected in Mom2B at the end of the first, second and third pregnancy trimester as well as at 6, 14, 24, 36 and 50 weeks postpartum.

The second self-report inventory was the five-item World Health Organization wellbeing index (WHO-5) (23, 24) that investigates the degree of subjective quality of life based on positive mood (good spirits, relaxation), vitality (being active and waking up fresh and rested), and general interest (being interested in things). Each item was rated from 0 to 5; the total score ranged from 0 to 25. According to the instructions, a percentage value was calculated by multiplying the score by 4 and thus obtaining a scale from 0 (worst) to 100 (best). A percentage score below 50 was interpreted as indicating risk of depression (23, 25).

The following questions concerning the impact of the pandemic were assessed three times during pregnancy between gestational weeks 11-20, 22-30 and 32-42, as well as during postpartum weeks 3-20 and 22-42:

1. *Have you received treatment for Covid-19?* (a)Yes; b) No)
2. *Have you had symptoms similar to the description of Covid-19?* (a)Yes, tested positive; b) Yes, tested negative; c) Yes, was not tested; d) No, but someone in my family had symptoms and e) No, and no one in my family had any symptoms)
3. *How is your life situation affected by the pandemic?* (a) It is not affected; b) Only slightly affected; c) There is a lot in my life that is affected and d) Almost everything in my life is affected)
4. *Which option describes your current situation?* (a) My situation is about normal; b) I am more isolated, and I am negatively affected; c) I am more isolated, but I feel OK and d) I am more isolated and this is mostly positive for me)

Confirmed or possible Covid-19 infection was defined as the participant replying with one of the following: 1a, 2a, 2b, 2c. Life situation was defined as “affected” when the participant replied with 3c or 3d and unaffected when replying with 3a or 3b. There was also an open question regarding the potential impact of the pandemic on the maternity and delivery health care received. Because the absolute majority of participants were pregnant during this analysis, only pregnancy assessed variables were included in this study.

### Mobility data

Movement data was collected using the GPS sensor on the phone. To save battery, the GPS sensor was iteratively turned on for 60 seconds and turned off for 10 minutes. A random offset between -100 km and +100 km was applied in both latitude and longitude to preserve the privacy of the participants. In total, GPS data was available for 1400 participants. For each participant, records before January 2020 and after October 31, 2020 were removed. Records with a high inaccuracy of the GPS sensor (>100 m) were also removed. Participants were also removed if they had less than 100 records remaining. After data cleaning, 1189 participants were included in this analysis.

Since the GPS sensor is turned on and off with a fixed interval, the absolute distances travelled for each user every month could not be calculated; instead, the relative difference in distance travelled away from their relative “home position” between months was used. The approximate home location of each participant was estimated by taking the median of the latitude and median of the longitude, assuming that each participant spends the majority of her time at home, and adding a radius of 100 meters. Next, for each GPS record outside the “home location”, the Euclidean distance to the home location was calculated. For each participant, the monthly median of the distance travelled away from home was used. The aggregated between-participant summary statistics were computed over these monthly medians.

### Google search volumes data

The interest of internet users for the situation around the pandemic over time (January-October 2020) in Sweden was assessed using Google Trends (https://trends.google.com/trends/explore?geo=SE&q=Corona). The numbers procured represent search volumes relative to the highest point on the chart (week with highest number of searches) for the given region and time. A value of 100 denotes the timepoint for the peak popularity for the term. A value of 50 means that the term is half as popular. A score of 0 means there was not enough data for this term.

### Statistical analyses of quantitative data

The EPDS, EPDS-3A and WHO-5 total monthly mean scores over time (from January to October 2020) were compared using the Analysis of Variance (ANOVA) regardless of the pregnancy time (early, middle or late). The distribution of the prevalence of positive screening for depressive symptoms (EPDS score over the cut-off of 12 points), anxiety symptoms (EPDS-3A score over the cut-off of 6), and low well-being (WHO-5 score below the cut-off of 50), over time were evaluated using the Pearson chi-square statistic. The associations between EPDS, EPDS-3A and WHO-5 in pregnancy and the self-reported impact of the pandemic during early (11-20 gestation weeks), middle (21-30 gestation weeks) and late (31-42 gestation weeks) pregnancy, were assessed using the Pearson’s chi-square test, student’s t-test and ANOVA as appropriate. The Spearman correlation coefficients and the respective *p*-values were calculated to assess possible correlations between the monthly mean total scores in EPDS, EPDS-3A, WHO-5 (total score and each subscale) and (1) the monthly median distance away from home, as well as with (2) google search volumes on “corona”.

All reported *p*-values were based on two-sided tests and the 5% significance level was used for hypothesis testing. Analyses were performed using the SPSS software, version 22.0.

### Qualitative data and analysis

Among questions on the mobile application regarding COVID-19, participants could comment in two open questions, on any impact on (1) their antenatal health care or birth plan, and (2) the delivery care. The questionnaire was sent out via the mobile app at three times during gestational weeks 11-22, 24-32 and 32-42. Analysis of the free text responses were performed using systematic text condensation (26) to comprehend the experiences of pregnant women of healthcare during Covid-19 pandemic. Every answer was divided into “meaning units” and the main content of the meaning units were identified and categorized.

## Results

The background characteristics of the 1345 women included in the Mom2B-study are presented in Table 1. The majority of participants were born in Sweden and were primiparous. Moreover, most women had university level education and lived with a partner. More than half of women reported having experienced depression earlier in life (Table 1).

**Table 1.**
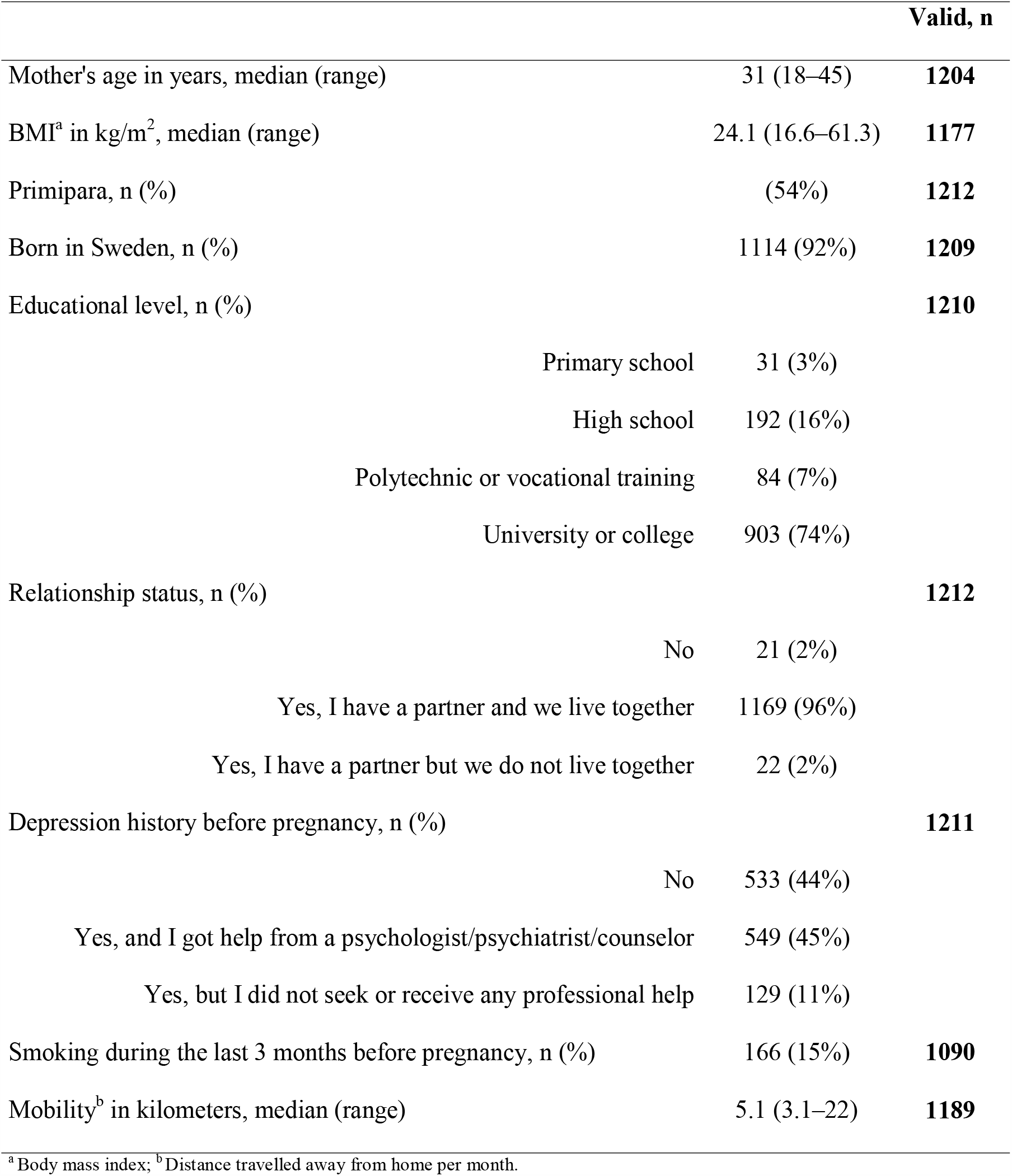
Background characteristics of the participating women in the Mom2B-study (n=1345)

Analyses of the mental health scores over time showed that the mean scores for depressive symptoms and anxiety peaked in April 2020 and increased again in October 2020 (Figure 1a), with 24-25% of pregnant women reporting EPDS scores over cut-off during April and May (Table 2). Statistically significant differences over this period were observed for EPDS and WHO-5 total scores using ANOVA (p=0.045 and p<0.001 respectively). The wellbeing scores were highest in January, as well as in June-August 2020. Mobility also peaked during the summer months of 2020 (Figure 1b). The google searches regarding the corona pandemic peaked in March, closely before the peak in symptoms of depression and anxiety, and started rising again in late October 2020 (Figure 1c). The prevalence of women scoring above cut-off in EPDS in 2020 was much higher, often more than double, in comparison with data from 2009-2019 (Table 2), when values were relatively stable across the seasons; no statistical testing was applied, as these data were derived from another cohort.

**Table 2.** Monthly distributions of prevalence for positive screening for depression, anxiety and low wellbeing

|  | Jan | Feb | Mar | Apr | May | Jun | Jul | Aug | Sep | Oct | *p*-value <sup>a</sup> |
| --- | --- | --- | --- | --- | --- | --- | --- | --- | --- | --- | --- |
| **Mom2B study (2020)** |  |  |  |  |  |  |  |  |  |  |  |
| Low wellbeing<sup>b</sup> (%) | 37 | 50 | 52 | 49 | 50 | 42 | 43 | 45 | 47 | 46 | <0.001 |
| Anxiety<sup>c</sup> (%) | 8 | 8 | 10 | 11 | 8 | 6 | 5 | 6 | 6 | 7 | 0.477 |
| Depression<sup>d</sup> (%) | 10 | 20 | 18 | 25 | 24 | 21 | 17 | 17 | 15 | 20 | 0.292 |
| **BASIC study (2010-2019)** |  |  |  |  |  |  |  |  |  |  |  |
| Depression<sup>d</sup> (%) | 8 | 9 | 10 | 10 | 10 | 9 | 10 | 10 | 10 | 8 | 0.779 |

<sup>a</sup> chi-square derived *p*-value<sup>b</sup> based on a WHO-5 total score < 50<sup>c</sup> based on an EPDS-3A total score > 6<sup>d</sup> based on an EPDS total score > 12

**Figure 1a.**
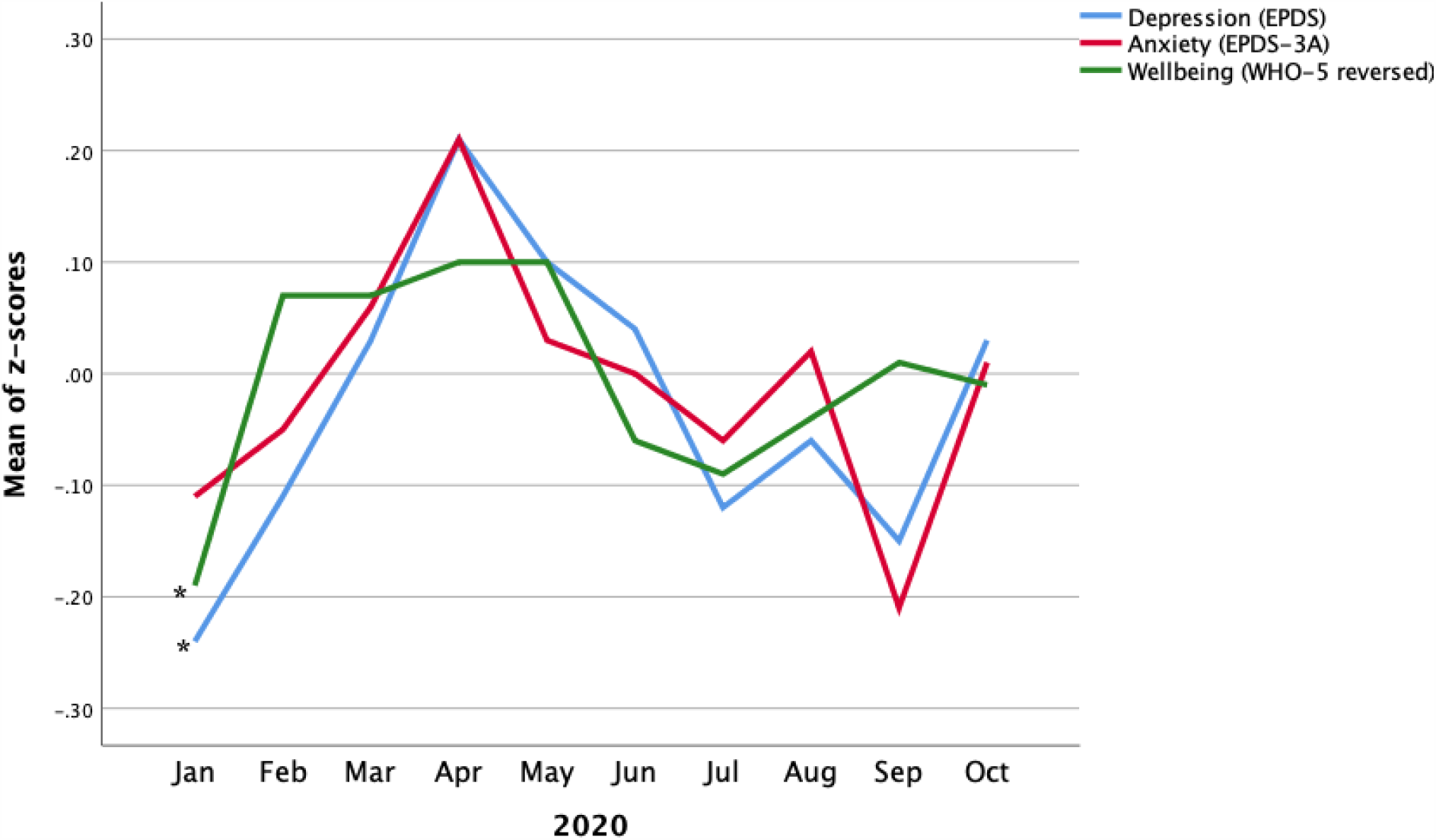
Mean of z-scores for monthly total scores of depression, anxiety and low wellbeing during January-October 2020

**Figure 1b.**
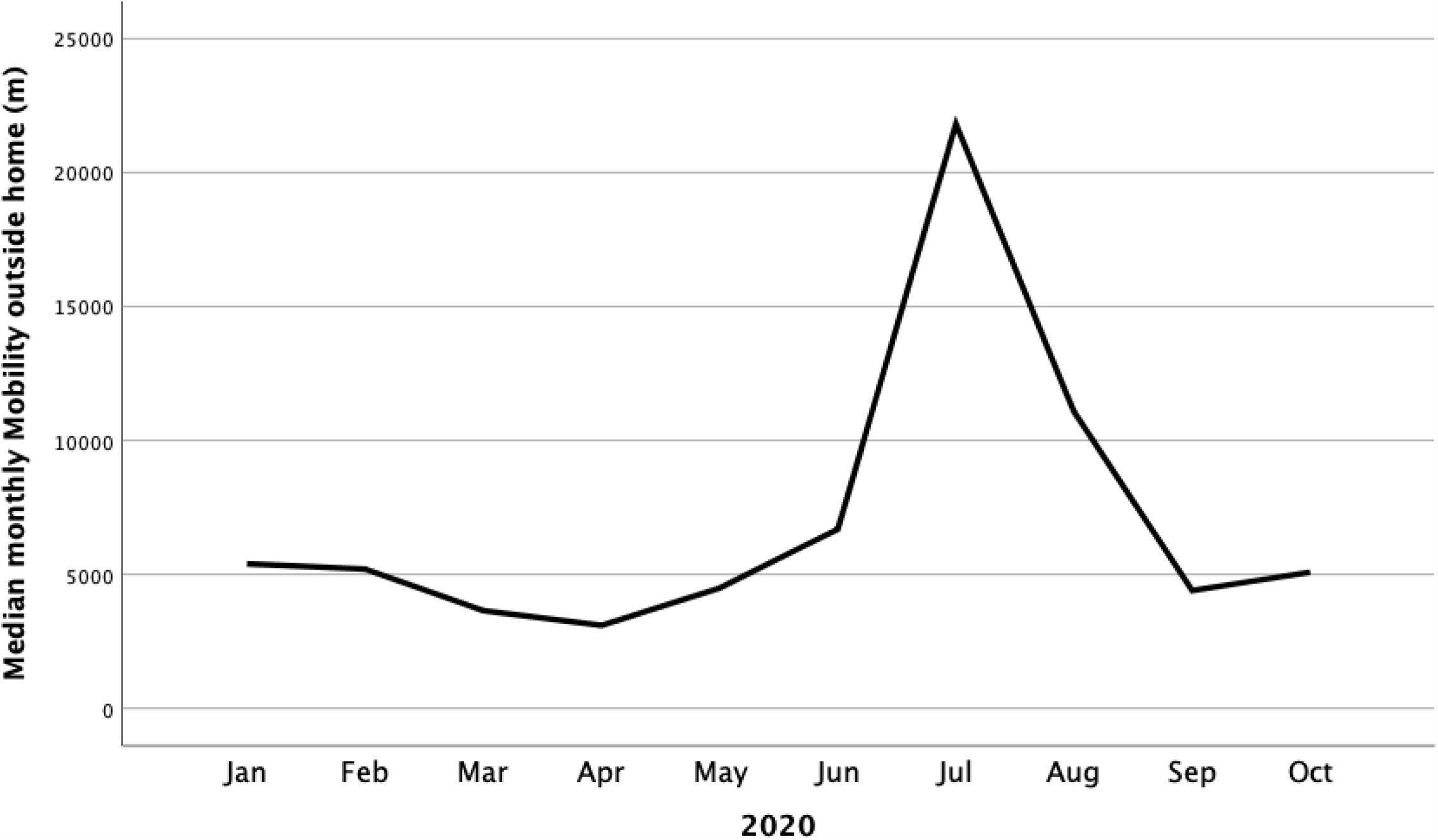
Median of monthly distance away from home (in meters) based on GPS data during January-October 2020

**Figure 1c.**
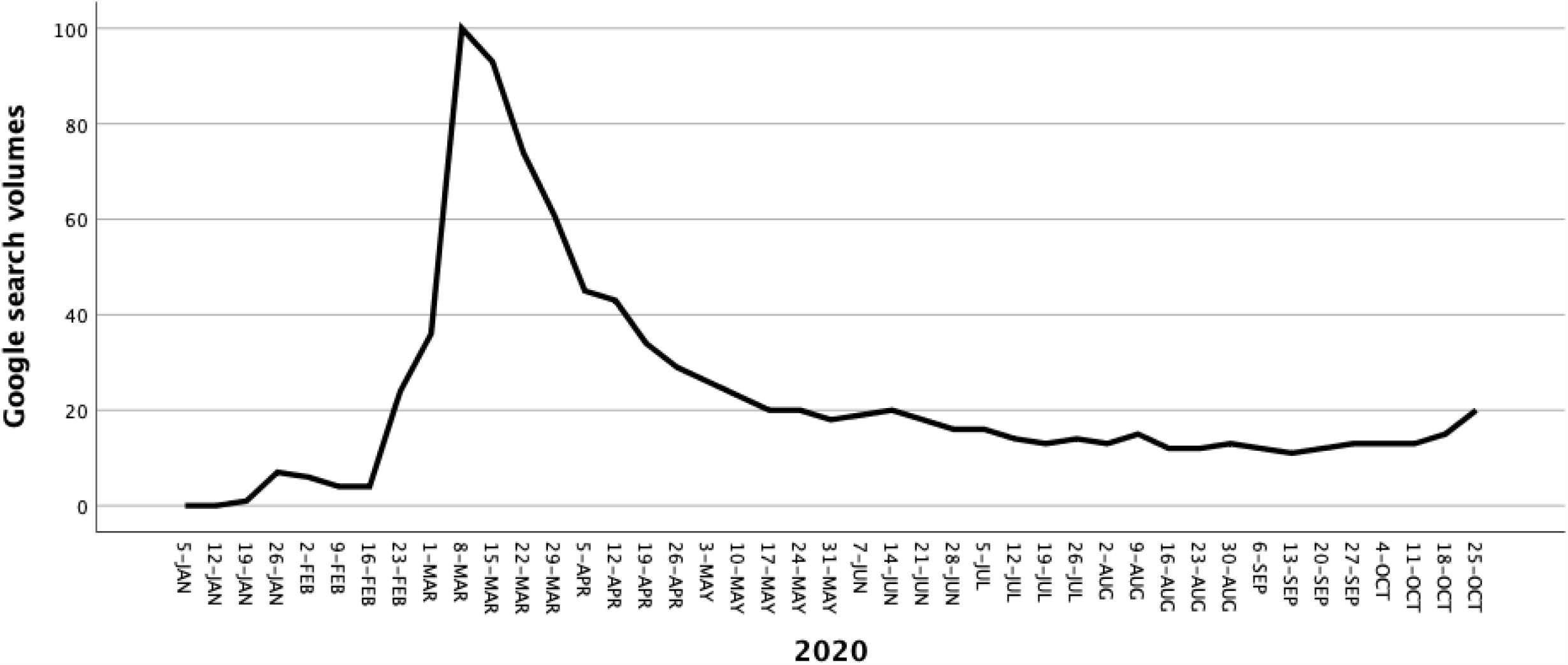
Google search volumes for “corona” in Sweden, during January-October 2020

The majority of participants reported that their life situation was only slightly affected by the COVID-19 pandemic (at any of the three-pregnancy time-points) (Table 5). Most women felt more socially isolated, and about one third of them perceived it as difficult (Table 5). Women reporting having no symptoms had higher anxiety symptoms (10%) in gestation week 22-30 than those with symptoms (3%) or a close friend or relative with symptoms (4%) (Table 3). Otherwise, no differences were observed among participants with Covid-19 related symptoms regarding depressive symptoms or well-being (Table 3).

**Table 3.**
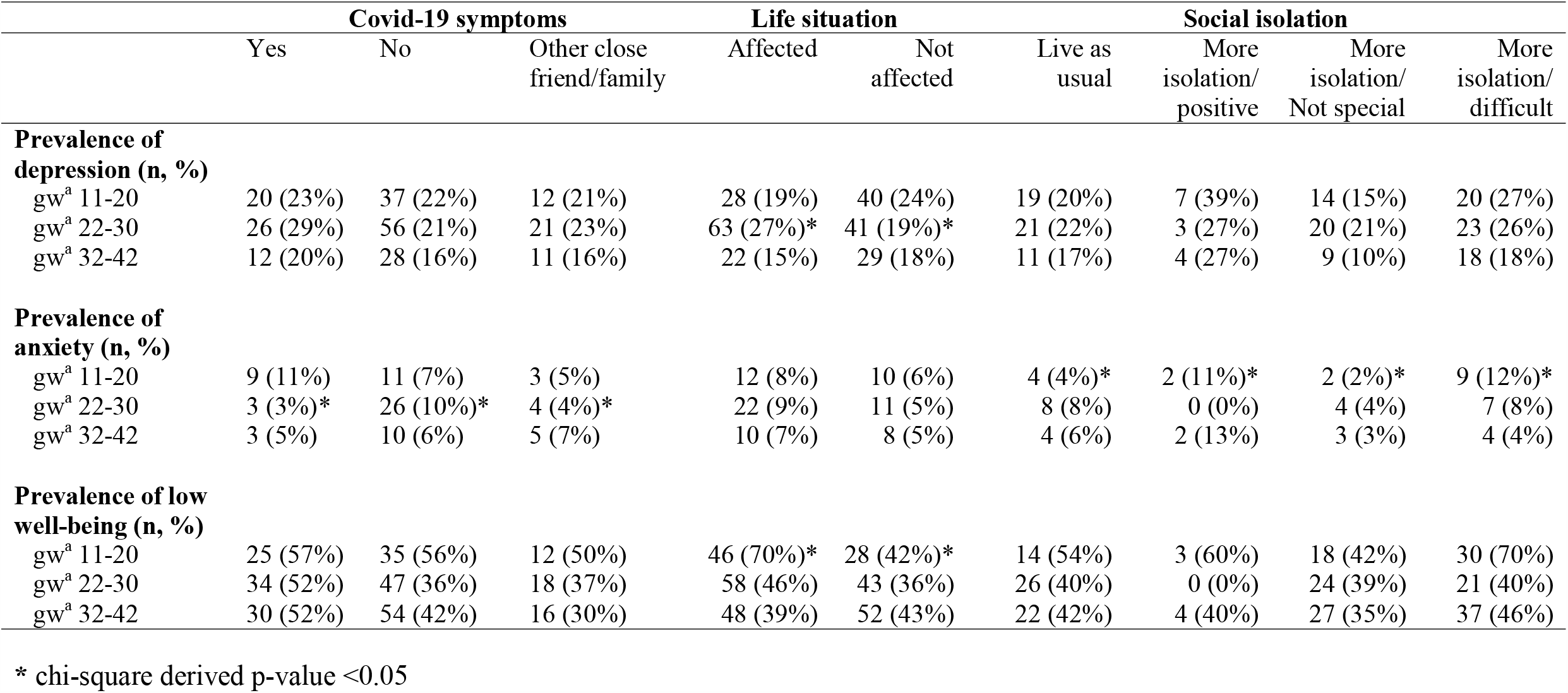
Prevalence of positive screening for depression, anxiety and wellbeing, by confirmed or possible Covid-19 infection and self-reported impact on life situation.

**Table 5.** Responses on questions concerning the COVID-19 pandemic

|  | Early<br>pregnancy<br>(gw<sup>a</sup> 11-20)<br>n=373 (%) | Middle<br>pregnancy<br>(gw<sup>a</sup> 22-30)<br>n= 543 (%) | Late<br>pregnancy<br>(gw<sup>a</sup> 32-42)<br>n=443 (%) |
| --- | --- | --- | --- |
| Have you had symptoms similar to the description<br>of Covid-19? |  |  |  |
| *Yes, but I have tested negative* | 12 (3%) | 14 (3%) | 11 (2%) |
| *No, not me and no one in my household or other*<br>*close friend/relative* | 193 (53%) | 305 (57%) | 237 (55%) |
| *No, not me but someone or more in my household or*<br>*another close friend/relative* | 70 (19%) | 109 (20%) | 95 (22%) |
| *Yes, but I have not been tested* | 81 (22%) | 101 (19%) | 85 (20%) |
| *Yes, and I have tested positive* | 9 (3%) | 7 (1%) | 5 (1%) |
| Many are more socially isolated during the<br>pandemic and this can be experienced in different<br>ways. Which option best suits your situation? |  |  |  |
| *I live about as usual* | 108 (32%) | 117 (33%) | 99 (25%) |
| *I'm more isolated, and it feels mostly difficult* | 95 (28%) | 99 (28%) | 134 (33%) |
| *I'm more isolated, but it does not feel special* | 113 (33%) | 120 (34%) | 147 (37%) |
| *I'm more isolated, and it feels mostly positive* | 22 (7%) | 15 (5%) | 21 (5%) |
| Life situation affected during the pandemic |  |  |  |
| *My life is not affected* | 21 (6%) | 13 (2%) | 23 (5%) |
| *I am only slightly affected* | 177 (47%) | 249 (46%) | 207 (47%) |
| *There's a lot in my life that is affected* | 144 (39%) | 233 (43%) | 164 (37%) |
| *Almost everything in my life is affected* | 30 (8%) | 48 (9%) | 46 (11%) |

<sup>a</sup> gw = gestational week

The proportion of participants with depressive symptoms over the cut-off during gestational week 22-30 was higher among those reporting that that their lives were affected by the pandemic (27% vs 19% in those not affected, *p*<0.05, Table 3). Similarly, the proportion of participants with lower wellbeing during gestational week 11-20 was higher among those reporting impact by the pandemic on their lives (70% vs 42% in those not affected, *p*<0.05, Table 3). No statistically significant differences were found in other periods in pregnancy or with regards to anxiety symptoms.

Furthermore, participants who reported they were socially isolated, more often reported anxiety scores over the cut-off (12 vs. 4%) during gestational weeks 11-20. Interestingly, among those reporting more social isolation, there was a higher prevalence of anxiety symptoms among those reporting that the isolation felt mostly hard (12%) or mostly positive (11%), compared to those reporting that it did not feel special (2%) or those reporting no social isolation (4%) (Table 3).

A positive correlation was found between the total score of wellbeing and mobility (rho=0.76, p=0.01), reaching a rho=0.83, p=0.003 for the WHO-5 question “My everyday life has been full of things that interest me” (Figure 2a). Similarly, strong statistically significant positive correlations were shown between both depression and anxiety scores and search volumes on corona via Google Trends (rho=0.87 and 0.83 respectively; Figure 2b). In a sensitivity analysis excluding the month of July, both the total score of wellbeing as well as the WHO-5 question “My everyday life has been full of things that interest me” remained statistically significantly correlated with mobility (rho=0.73, p=0.03 and rho=0.82, p=0.007 respectively).

**Figure 2a.**
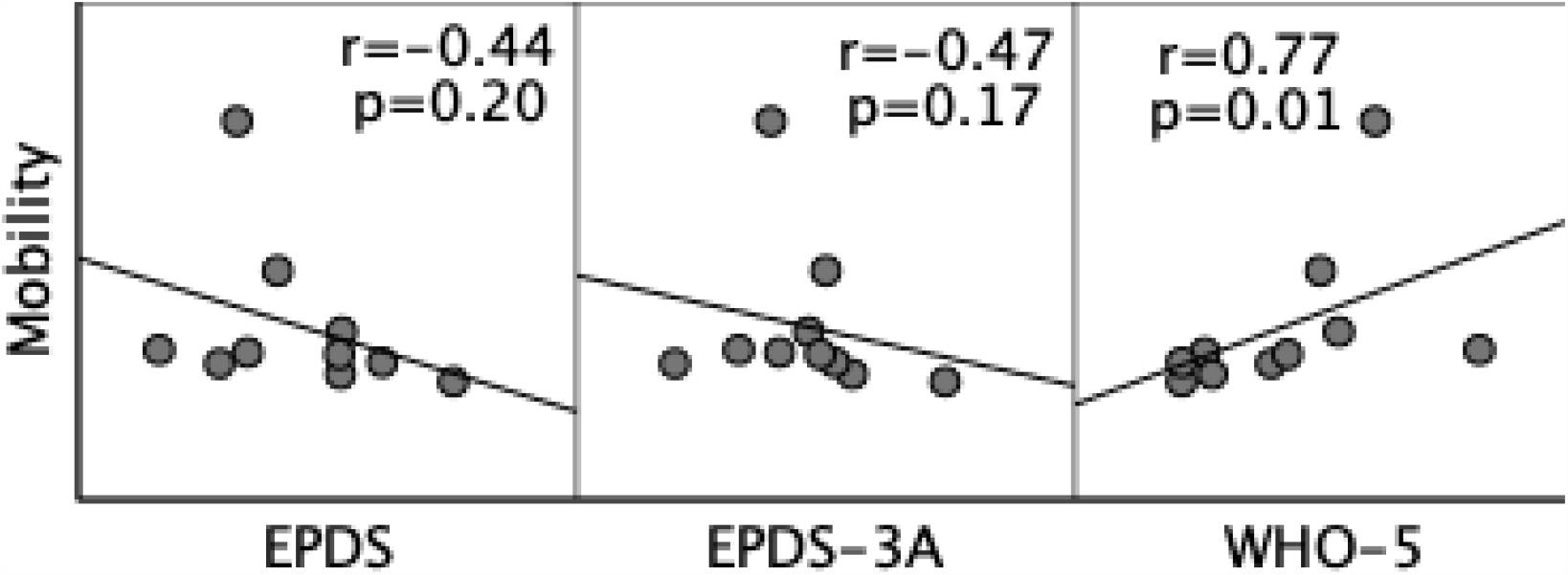
Scatterplots for the monthly mean in total scores in EPDS (depression), EPDS-3A (anxiety) and WHO-5 (wellbeing) by mobility (monthly median distance away from home).

**Figure 2b.**
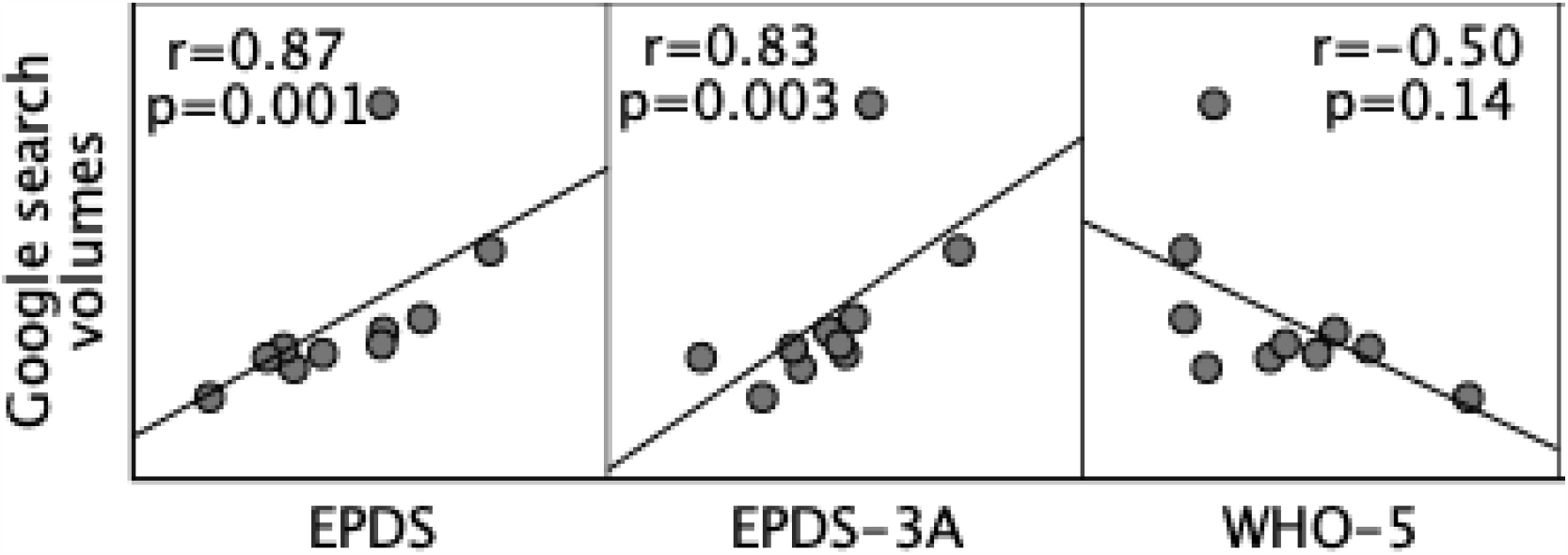
Scatterplots for the monthly mean in total scores in EPDS (depression), EPDS-3A (anxiety) and WHO-5 (wellbeing) by Google search volumes on “corona”.

Table 4 presents a compilation of the women’s free responses to the open questions regarding healthcare during the pandemic across pregnancy and childbirth. The answers have been divided into different categories, and next to it, one or more quotes. The participants expressed loss of support when partners were excluded from attending visits at the maternity center and ultrasound examinations. Many comments addressed the risks of the partner missing out during the delivery of the child, as no-one with the slightest symptoms could be accompanying the delivering woman. Moreover, other support stemming from relatives or doulas was not allowed, which further enhanced worry and distress. When participants mentioned that check-ups had been canceled/postponed, they usually referred to ultrasound examinations or combined ultrasound and biochemical screening. Moreover, they also reported meetings with other professionals, such as social workers postponed or cancelled. Some participants also felt unsafe about their health care. They experienced miscommunication and insufficient information from health care professionals. Referrals had been rejected due to the strain caused by COVID-19. Some comments referred to the notion that online or phone meetings were not satisfactory regarding the level of care.

**Table 4.** Categorization of participants’ experiences of how maternity care was affected during the pandemic.

| **Categories** | **Quotations for illustration** |
| --- | --- |
| ***The quality of care***<br>- **Lack of information**<br>- **Shortage of staff**<br>- **Inferior treatment**<br>- **Fewer physical examinations**<br>- **Postponed care** | "I don't feel safe with my health care!"<br>"Appointments over the phone do not provide as good care "<br>"Due to restrictions about physical appointments, I am not sure that I got the correct diagnose." |
| ***Childbirth***<br>- **Reduced support due to visitor restriction policies**<br>- **Fewer possibilities to choose type of care**<br>- **Feelings of uncertainty** | "I now plan to give birth at home with a midwife"<br>"My partner may not be able to attend the childbirth at all if the slightest cold symptoms occur, so I must have back-up I will probably have to stay [without my partner] in the maternity ward, something I feel very anxious about" |
| ***Maternity care***<br>- **Reduced support for fear of childbirth/ due to visit restrictions**<br>- **Cancelled/postponed visits**<br>- **Preparatory courses cancelled**<br>- **New local routines**<br>- **Concern for Covid-19 infection** | "The early ultrasound was cancelled"<br>"Very sad and worried, cried a lot when I found out that my partner was not allowed to come to [the test]. Made me feel really bad"<br>"Very frustrated and worried about the restriction policy, which I experience as arbitrary and incomprehensible" |
| ***Seeks health care to a lesser extent*** | "I do not seek health care "unnecessarily/ for non-acute problems"<br>"I was advised not to seek care" |
| ***Social relationships***<br>- **Parent meeting and new contacts** | "I worry about not being able to meet other mothers as I did with my first child" |
| ***No impact on health care*** | "All my appointments have been completed" |

## Discussion

### Main findings

We used a novel study method, the research mobile application Mom2B, to assess, for the first time, in nearly real time self-reported perinatal depressive symptoms and data on SARS-CoV-2 treatment, symptoms, impact on everyday life and perceived health care, as well as symptoms of depression, anxiety and wellbeing. Data from 1345 pregnant women at a national level between January and October 2020 in Sweden were utilized to investigate the potential changes in mood and behaviors during the pandemic. An increase in depressive symptoms and decrease in total wellbeing was shown during the months with the highest pandemic impact in Sweden (April, May) followed by normalization during the subsequent months and a new peak in October, when cases started to surge again. Approximately 24% of pregnant women reported depression scores above cut-off in screening instruments during April and May, with a subsequent tendency to increase again as the infectious rates started rising by October 2020. These results are in sharp contrast to the reported rates, estimated at around 12%, in different Swedish cohorts during earlier years (27, 28). No associations were found between treatment or symptoms of SARS-CoV-2 and changes in mood and general wellbeing.

### Previous literature

The present findings are in general congruent with recent research providing mounting evidence for an increase in the levels of anxiety and depression among pregnant women during the COVID-19 pandemic. Several studies in China, Japan, Turkey, Pakistan, Qatar, US and Canada have reported that 15% up to 35% of women scored high EPDS scores (above 12-13) during pregnancy and early postpartum period because of the SARS-CoV-2 pandemic (2, 6, 7, 14, 29-35). A Canadian study also showed higher levels of depressive and anxiety symptoms, dissociative symptoms, symptoms of post-traumatic stress disorder, negative affectivity and less positive affectivity in a COVID-19 cohort compared to a pre-COVID-19 cohort of pregnant women (36). Analyzing the comments from pregnant women in the Mom2B cohort, it is noteworthy that most women reported less access to healthcare facilities, less satisfaction about the quality of healthcare delivery, as well as less support during pregnancy; in general, most emotions and experiences were negative. However, not all women provided comments and thus, we cannot conclude whether the expressed emotions were representative of all women; however, the present results are in line with recent studies. Specifically, consistent with our results, a previous Swedish study including a regional population-based cohort showed increased levels of health-related worry in pregnant women during the months of the pandemic (February-August 2020) (37), whereas other studies have also reported increased levels of anxiety and depression due to loss of supportive perinatal services and uncertainty about the novel coronavirus disease (38).

At baseline, Sweden provides healthcare at minimal cost to all inhabitants including mental health services (39, 40). Moreover, Sweden is among the countries with the lowest rates of preterm births (between 5-6%) (41), likely due to the economic stability of the country and the focus of the government on perinatal health, as well as on the timely and adequate access to healthcare services (42). Despite the fact that the Swedish COVID-19 strategy did not include a general lockdown, but was instead based on voluntary social distancing guidelines (16), we still find a clear association of theCOVID-19 pandemic with the mental health of pregnant women. Notably during spring, even in Sweden, some extra screenings for chromosomal aberrations and services for fear of childbirth were not available in some regions; and concurrently, partners were not allowed to stay after delivery at the hospital. During the whole period, healthcare has been affected through reduced staff, fewer physical appointments and restrictions to access healthcare for individuals with symptoms of common cold. While online consultations are an effective way of providing safe care during the outbreak, the basic elements of the patient-doctor or midwife relation may be reduced, and the sense of genuine connection was lost (43), which was also reported by the participants in the Mom2B study. Further research is needed to understand individuals’ thoughts on access to care in different health care systems during the pandemic.

It is of note that women who did not report symptoms of the COVID-19 infection self-reported higher levels of anxiety. In general, data from the Mom2B cohort indicated that infected women did not feel greater anxiety and depression compared to not affected women. One reason might be that most infected cases were mild; thus, the SARS-CoV-2 mild infection might be a relief to these women compared to the worry of the otherwise unknown, and potentially severe impact of the disease. Moreover, although data on other adverse effects of COVID-19 notably on the rates of preterm birth and other obstetric complications is quickly being gathered and analyzed, findings remain inconsistent so far; therefore, women in Sweden did not receive any relevant information on these complications during their visits to maternity care.

Since the beginning of the pandemic, pregnant women in most countries have faced the effects of social isolation and restrictions in mobility, e.g., in Ireland, an exercise limit of one to two kilometers from home, while all other journeys outside to be made only for provision of essential services and goods (44). Previous studies have reported a significant association of social isolation with exacerbation of domestic violence and poor mental health outcomes (44-46). A Spanish internet-based study showed that confinement decreased both the level of vigorous and moderate physical activity that pregnant women had engaged in, as well as the time they spent walking, and doubled the number of hours they spent sitting; moreover, confinement negatively affected their health-related quality of life (47). Our results showed that not only social isolation, but also the emotional connotations of the fewer health care appointments and other downstream effects of the pandemic are probably associated with increased risk of anxious symptoms. They further support the findings linked to restricted movement, showing a strong inverse correlation between the distance travelled away from home (reduced during April-May, and again in October, according to voluntary recommendations) and wellbeing. The directionality of the association is difficult to decipher using group data; yet, future studies are expected to examine if movement changes precede the decrease in wellbeing. In addition, the possible role of vacations during summer months, with low cases and increased mobility for the holidays might contribute to these findings. Nevertheless, earlier studies did not in general show statistically significantly lower levels of perinatal depression symptoms during summer months compared to spring (28). Further, sensitivity analysis excluding July, which is the usual vacation month for Sweden, did not significantly alter the results.

Health-seeking behaviors, such as online searches, have been extensively analyzed for monitoring other diseases, such as seasonal influenza in the past (48); however, to our knowledge, such strong correlation, as the one found herein between google search volumes for an infectious disease (such as COVID-19) and mental health outcomes, has never been reported before. If this finding is replicated in future research, internet search volumes for global health emergencies could be a useful proxy of the mental health status at population level, enhancing public health monitoring and interventions.

### Strengths and limitations

The novel way provided through the Mom2B application to introduce new questions and gather self-reported information in a user-friendly way at national level is a strength of this study. Future research could further address whether possible preventive interventions might be incorporated and delivered directly through the application, adding to the health care systems arsenal during crises.

Among the limitations of the present study, we should first acknowledge that selection bias should be considered: only 1% of the background eligible population contributed data; the application cannot be used by women not able to communicate in Swedish, or those not owning a mobile smartphone (8%). Nonetheless, the present results are in line with another recent Swedish study with better representation (37). We cannot exclude a potential bias in the results due to the self-reported nature of the information gathered. In addition, we used the EPDS and WHO-5 scores for outcome assessment, which are validated screening questionnaires, and sensitive in identifying women likely to suffer from perinatal mood or anxiety disorders; however, these tools are not sufficient to establish the diagnosis of perinatal depression or anxiety. Data on mobility at the individual level would have allowed for more advanced analyses, which could address directionality in the associations with mood; this will be possible in the near future. Lastly, the ongoing use of Mom2B, allowing for a longer follow-up period and the ongoing recruitment of new participants will allow further analyses to be performed, controlling for potential confounders and mediators, and looking even to other passively collected data on mobile and internet use, which were not feasible at this stage.

## Conclusions

Utilizing a novel application, we observed an increase in depressive symptoms and decrease in wellbeing during the first months of the pandemic in Sweden, followed by a return to baseline levels during summer, and a new rise in October following the new surge in COVID-19 cases in the country. Strong correlations between mobility as well as internet searches and wellbeing were noted. Key themes were identified including worry about the impact of the virus on health, worry about access to healthcare, as well as social and economic impacts secondary to the pandemic. The Mom2B application further allowed us to gather data in a user-friendly way among pregnant women as the pandemic was evolving; such tools are expected to be useful as other disasters occur and unfold. Lastly, the present results, supported by previous studies in different settings, suggest that there may be protective and resilience factors associated with different healthcare systems, highlighting the need to consider the policy decisions’ impact on perinatal mental health. As the goal of maternity care is to identify and reduce the risks of ill health, mental ill health should be promptly identified, especially during periods of crises, whereas substantial preventive efforts should be put in place to minimizing the adverse events on perinatal mental health.

## Data Availability

Data is avaliable upon reasonable request.

## Acknowledgements

We would like to thank Oskar Burman, Catherine Axfors, Edith Ngai and Diem Nguyen for their invaluable help with the Mom2B infrastructure.

